# Brugada ECG detection with self-supervised VICReg pre-training: a novel deep learning approach for rare cardiac diseases

**DOI:** 10.1101/2024.03.29.24305072

**Authors:** Robert Ronan, Constantine Tarabanis, Larry Chinitz, Lior Jankelson

## Abstract

Existing deep learning algorithms for electrocardiogram (ECG) classification rely on supervised training approaches requiring large volumes of reliably labeled data. This limits their applicability to rare cardiac diseases like Brugada syndrome (BrS), often lacking accurately labeled ECG examples. To address labeled data constraints and the resulting limitations of supervised training approaches, we developed a novel deep learning model for BrS ECG classification using the Variance-Invariance-Covariance Regularization (VICReg) architecture for self-supervised pre-training. The VICReg model outperformed a state-of-the-art neural network in all calculated metrics, achieving an area under the receiver operating and precision-recall curves of 0.88 and 0.82, respectively. We used the VICReg model to identify missed BrS cases and hence refine the previously underestimated institutional BrS prevalence and patient outcomes. Our results provide a novel approach to rare cardiac disease identification and challenge existing BrS prevalence estimates offering a framework for other rare cardiac conditions.

## Introduction

Deep learning algorithms, like convolutional neural networks (CNN), have been adapted for analyzing 12-lead electrocardiograms (ECGs)^1^. The resulting artificial intelligence (AI) models can offer equivalent or higher diagnostic accuracy compared to expert or rule-based computer interpretations^2,3^. These traditionally supervised machine learning approaches to ECG algorithmic diagnosis require large volumes of positive label ECGs often numbering in the thousands^1^. This limits their generalizability to rare cardiac diseases and more broadly to small institutional datasets of common cardiac conditions. Brugada syndrome (BrS) is a rare condition associated with familial aggregation and premature sudden cardiac death exhibiting a coved-type or type 1 Brugada pattern in the right precordial ECG leads^4^. To overcome the limitations of supervised AI approaches in the diagnosis of rare cardiac diseases like BrS, we developed a novel deep learning model for BrS ECG classification using the Variance-Invariance-Covariance Regularization (VICReg)^5^ architecture for self-supervised learning, implemented as pre-training. We illustrate its superior performance compared to a neural network model without VICReg pre-training and use it to refine our institutional BrS prevalence.

## Results

### Model training

Within our institutional ECG database, we identified all patients with at least one MUSE-labeled BrS ECG (**Fig. 1**) and excluded them from training. The remaining patients without known BrS ECGs (Pre-training set) totaling 1,212,209 were split into the VICReg Pre-training, used to pre-train a base model, and Hold-out sets. In the initial, MUSE BrS positive arm, all ECGs belonging to patients with at least one MUSE-labeled BrS ECG were reviewed by an expert electrophysiologist (EP) yielding 123 and 214 patients with and without a BrS type 1 ECG pattern, respectively. In turn, these patients were divided into training and test sets. The resulting BrS Training set was used to train a state-of-the-art CNN model (hereafter called the Standard model), which was compared to a VICReg baseline model finetuned with the same BrS Training set.

**Figure 1:**
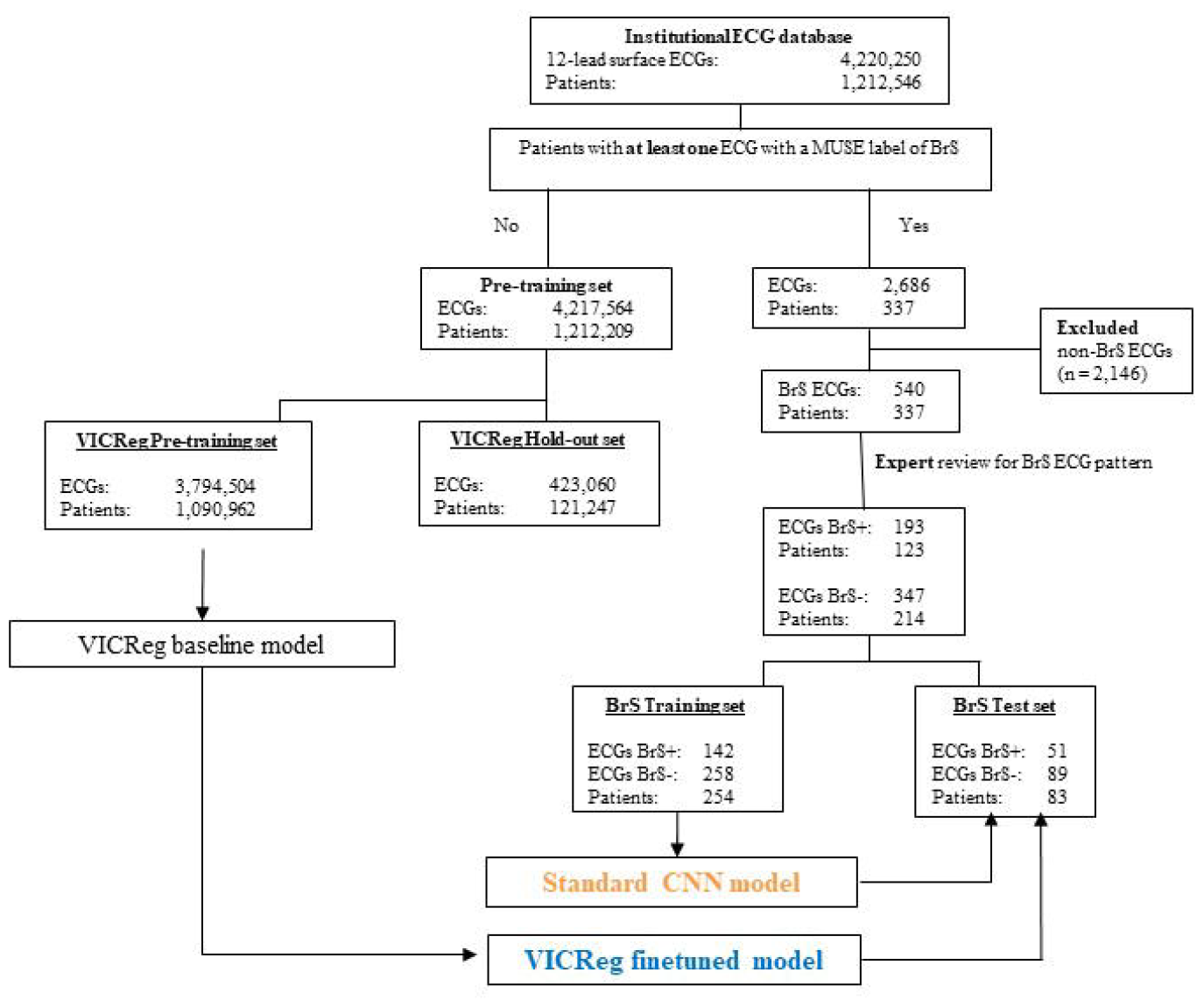
The study design flow chart for identifying and confirming patients with at least one type 1 BrS ECG, as well as for defining the training and test sets used for the Standard and VICReg model development. Abbreviations: BrS, Brugada syndrome; ECG, electrocardiogram.

### Model performance

The BrS Test set was used to evaluate model performance. The VICReg finetuned model outperformed the Standard model on all calculated metrics. Compared to the Standard model, the VICReg model had a greater area under both the receiver operating characteristic (**Fig. 2a**; AUROC 0.88 vs. 0.76) and precision-recall curves (**Fig. 2b**; AUPRC 0.82 vs. 0.67). The VICReg model also had superior performance in all remaining metrics including specificity (0.80 vs. 0.70), sensitivity (0.61 vs. 0.33), positive (0.76 vs. 0.65) and negative (0.80 vs. 0.70) predictive values, accuracy (0.79 vs. 0.69), Matthew’s correlation coefficient (0.52 vs. 0.29) and F1 score (0.67 vs. 0.44; **Table 1**). In turn, we applied the VICReg finetuned model to the VICReg Hold-out set discovering 34 new patients with BrS ECGs previously lacking the appropriate label. This new diagnosis was confirmed following ECG review by three EP physicians. We present an example type 1 BrS ECG in the VICReg Hold-out set, missed by the Standard but identified by the VICReg model (**Fig. 3 a, b**). Compared to age- and sex-matched controls, new BrS patients identified by the VICReg finetuned model had higher rates of atrial fibrillation (odds ratio 2.7 [1.2-6.0]), cardiac arrest (53.1 [9.4-300.1]), ventricular tachycardia (14.5 [5.3-39.3]), as well as implantable cardioverter-defibrillator (ICD; 11.8 [4.0-35.1]) and permanent pacemaker (PPM; 3.5 [1.1-11.7]) implantation (**Table 2**). Importantly, both the new and existing MUSE-labeled, EP-confirmed BrS patients had similarly increased rates of ICD and PPM implantation compared to age- and sex-matched controls (**Table 2**).

**Table 1.** Model performance metrics for the VICReg and Standard models.

| Model | VICReg | Standard |
| --- | --- | --- |
| AUROC | 0.88 | 0.76 |
| AUPRC/Average precision | 0.82 | 0.67 |
| Specificity | 0.80 | 0.70 |
| Recall/Sensitivity | 0.61 | 0.33 |
| Precision/Positive predictive value | 0.76 | 0.65 |
| Negative predictive value | 0.80 | 0.70 |
| Matthews Correlation Coefficient | 0.52 | 0.29 |
| Accuracy | 0.79 | 0.69 |
| Balanced Accuracy | 0.75 | 0.62 |
| F1 Score | 0.67 | 0.44 |

Abbreviations: AUROC, area under the receiver operating characteristic; AUPRC, area under the precision recall curve.

**Table 2.** Odds ratios for new BrS patients identified by the VICReg model (n=34) and existing MUSE-labeled, EP-confirmed BrS patients (n=123) compared to age- and sex-matched controls for a variety of cardiovascular conditions.

|  | New BrS patients identified by the<br>VICReg model (n=34) |  | Existing MUSE-labeled, EP-confirmed<br>BrS patients (n=123) |  |
| --- | --- | --- | --- | --- |
|  | OR (95% CI) | *p* value | OR (95% CI) | *p* value |
| Atrial fibrillation | 2.7 (1.2 - 6.0) | 0.021 | 0.7 (0.3 - 1.7) | 0.500 |
| History of cardiac arrest | 53.1 (9.4 - 300.1) | 0.004 | 4.6 (0.6 - 34.2) | 0.105 |
| Implantable cardioverter-defibrillator | 11.8 (4.0 - 35.1) | 0.014 | 12.2 (7.0 - 21.5) | <0.001 |
| Permanent pacemaker | 3.5 (1.1 - 11.7) | 0.043 | 3.3 (1.5 - 7.3) | 0.003 |
| Supraventricular tachycardia | 2.8 (0.7 - 12.1) | 0.168 | 0.5 (0.1 - 3.3) | 0.476 |
| Syncope | 2.3 (0.9 - 6.1) | 0.083 | 2.5 (1.5 - 4.2) | 0.002 |
| Ventricular tachycardia | 14.5 (5.3 - 39.3) | 0.008 | 0.7 (0.1 - 5.0) | 0.777 |

Abbreviations: BrS, Brugada syndrome; CI, confidence intervals; EP, electrophysiologist; OR, odds ratio.

**Figure 2:**
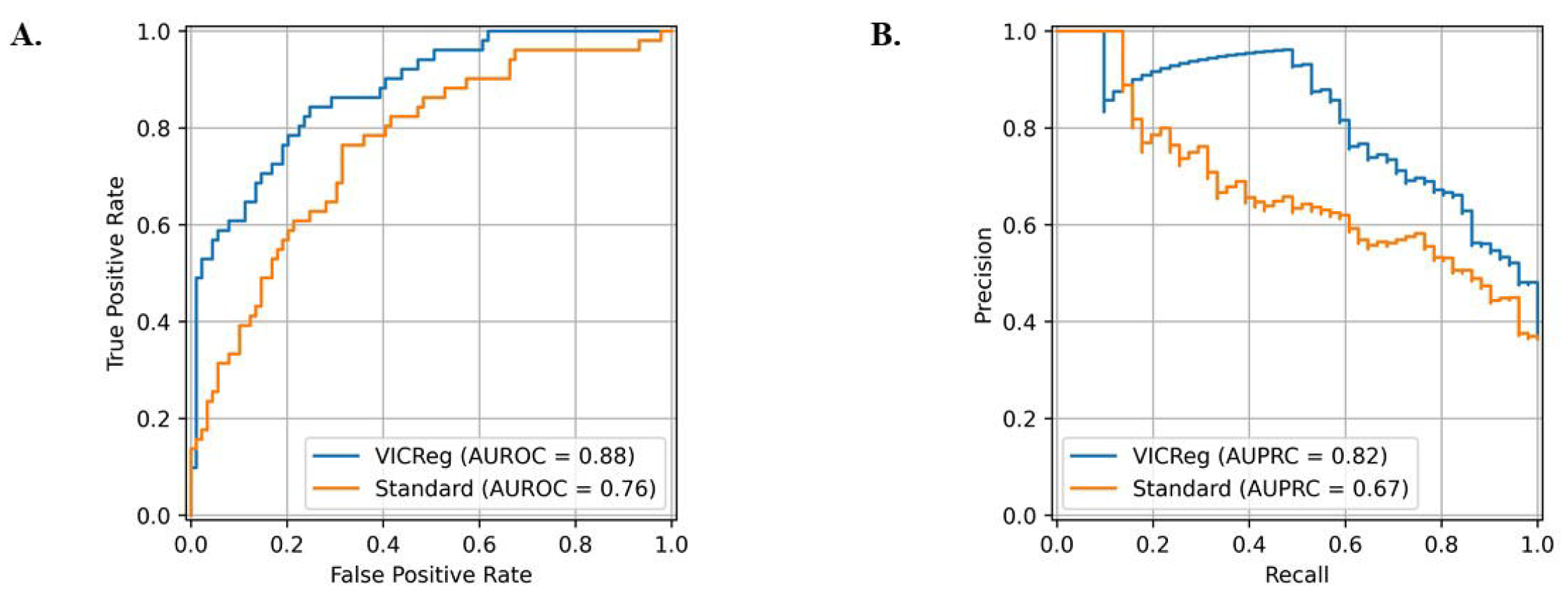
**A**. The receiver operating characteristic and **B**. precision-recall curves derived from the VICReg and Standard models.

**Figure 3:**
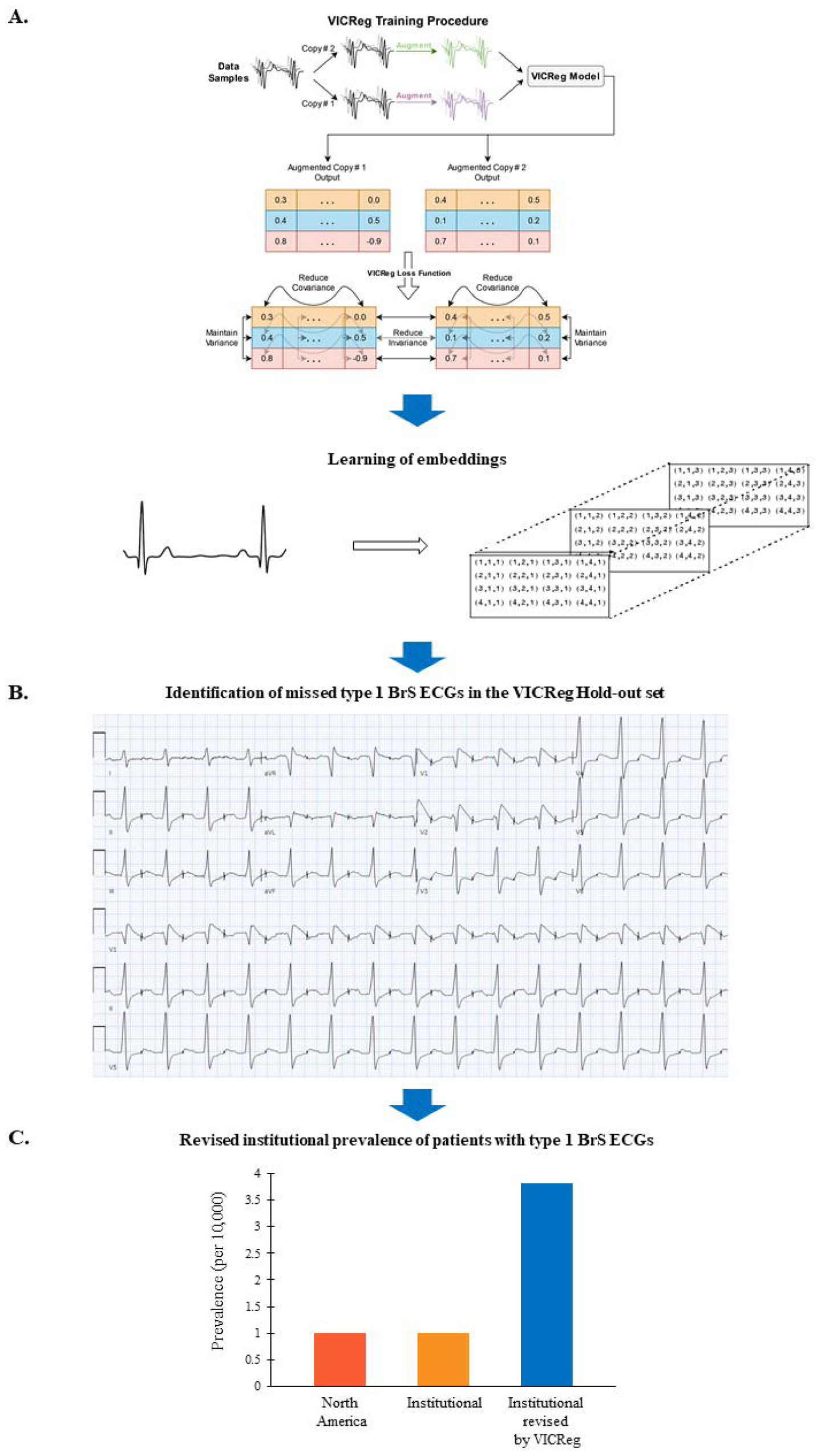
**A**. A graphical illustration of the VICReg pre-training approach for the learning of embeddings from the ECG signal. **B**. Example of a type 1 BrS ECG in the VICReg Hold-out set (i.e., without a MUSE BrS label) missed by the Standard but identified by the VICReg model. **C**. Prevalence of patients with type 1 BrS ECGs in North America per prior literature reports compared to our institutional prevalence before and after the application of the VICReg model. Abbreviations: BrS, Brugada syndrome; ECG, electrocardiogram.

## Discussion

In rare cardiac diseases there is a dearth of accurately labeled ECG examples. The rare nature of such conditions results in a lack of clinician familiarity with their ECG patterns and hence frequent inaccurate ECG labeling^6,7^ as evidenced by the reclassification of less than half of the MUSE-labeled ECGs as true BrS type 1 ECGs (**Fig. 1**). This challenge is further exacerbated by the poor performance of rule-based diagnostic algorithms^8^. Consequently, typical neural networks like the Standard model have limited diagnostic performance (**Fig. 2, Table 1**) requiring large volumes of reliably labeled training data. Pretraining with a self-supervised approach like VICReg addresses labeled data constraints in rare cardiac diseases, enabling the learning of embeddings (vectors) without requiring abundant examples or contrastive samples^5^ (**Fig. 3a**). With VICReg, a batch of samples is copied twice, and each copy is augmented separately. The two augmented batches are fed through two branches of a joint embedding architecture, which outputs two embedding (representation) matrices, consisting of embedding vectors for each sample in the batch. The VICReg loss function is computed on these embeddings and is designed to simultaneously reduce the distance between embedding vectors for each augmented copy of a sample (the invariance), maintain the variance above a threshold across the batch for each variable in the embedding, and reduce the covariance across the batch between pairs of variables in the embedding to zero^5^ (**Fig. 3a**). The VICReg model represents the first published ECG application of the VICReg architecture and holds generalization potential to other rare cardiac diseases like Long QT, Early Repolarization Syndrome, Left Ventricular Noncompaction and Arrhythmogenic Right Ventricular Dysplasia, among others. We illustrate the superior performance of this approach over “traditional” CNN models in a small dataset (n<200) of BrS ECGs (**Table 1**). Importantly, the VICReg model exhibited both high AUROC and AUPRC, with the latter being a key metric in imbalanced datasets and yet lacking in prior attempts^9–11^ at generating deep learning models for BrS ECG classification.

The lack of accurate ECG algorithms to reliably label rare patterns such as BrS, challenge our estimations of rare cardiac disease prevalence, as they rely on likely incomplete or erroneous datasets. Consequently, erroneous disease prevalence estimations reflect an inaccurate risk assessment. Indeed, the 34 new BrS patients identified by the VICReg model in the Hold-out set, alone increase our institutional BrS prevalence from 1.0:10,000 (95% CI: 0.8-1.2) to 1.3:10,000. Extrapolating from our findings in the VICReg Hold-out set, which is a random 10% sample of the Pre-training set that excluded patients with MUSE-labeled BrS ECGs, we could reasonably expect a total of 340 missed BrS patients in our entire institutional dataset. A revised estimation yields an institutional prevalence of 3.8:10,000 (95% CI: 3.5-4.2), approximately four times higher than previous North American BrS estimates of 0.5-1:10,000^4,12,13^ (**Fig. 3c**). This suggests that BrS is significantly underdiagnosed possibly owing to physicians’ lack of familiarity with its ECG findings and the poor performance of traditional automated diagnostic algorithms. The novel application of the VICReg self-supervised pre-training architecture could help overcome this hurdle bringing more BrS patients to clinical attention, sooner. Importantly, new BrS patients identified by the VICReg model appear to represent a clinically distinct patient population with a higher arrhythmic burden, as evidenced by higher rates of atrial fibrillation, cardiac arrest and ventricular tachycardia compared to age- and sex-matched controls (**Table 2**). In contrast, existing MUSE-labeled, EP-confirmed BrS patients did not have a significantly increased arrhythmic burden over age- and sex-matched controls yet shared the same increased odds ratios for ICD and PPM implantation (**Table 2**). This could reflect their timely BrS diagnosis, close monitoring and early intervention after earlier recognition of a type 1 BrS ECG pattern, as opposed to the missed cases now newly identified by the VICReg model.

In conclusion, we report a novel, VICReg pre-trained/ConvNext architecture that relies on the ECG signal alone to identify type 1 BrS ECGs. We demonstrate key advantages of the VICReg self-supervised pre-training approach in rare cardiac diseases through its superior BrS diagnostic performance compared to a neural network of equivalent architecture but without pre-training. Given the study’s single-center, retrospective design, future research should focus on the external validation and prospective application of the VICReg model. Our findings suggest that the model is poised to identify new and otherwise missed BrS cases and in the process redefine our epidemiological understanding of this cardiac condition, while providing a framework for other rare cardiac conditions.

## Methods

### Cohort selection

We identified all patients at our institution with at least one 10-second, 12-lead, surface, resting ECG between January 1, 1997 and December 31, 2021. The ECG waveforms were obtained from leads I, II, and V_1_-V_6_ and ECGs with a sampling rate of 250 Hz were up-sampled to 500 Hz. For each lead, we computed the mean and standard deviation of that lead’s values across the VICReg training set ECGs. These values were then used to normalize every ECG involved in model training and testing by subtracting the associated mean and dividing by the associated standard deviation. All patients with a mention of “Brugada” in their MUSE (GE Healthcare, Chicago, IL) ECG label were identified and separated (**Fig. 1**). An electrophysiologist reviewed these MUSE-labeled BrS ECGs confirming or rejecting a type 1 BrS ECG pattern, and their determinations were considered the ground truth. Patients with MUSE-labeled, expert-reviewed BrS ECGs were split into a 75% training and validation set (BrS Training set) and 25% test set (BrS Test set). Patients from the original full cohort lacking at least one MUSE-labeled BrS ECG were included in the Pre-training set. In turn, this was split into a 90% training and validation set (VICReg Training set) and 10% holdout set (VICReg Holdout set; **Fig. 1**). This study was approved by the hospital’s Institutional Review Board.

### Model Architecture

We used a modified version of the ConvNext^14^ family of CNNs as the basis for all our described models, adjusting the model architecture to utilize 1D instead of 2D convolutions (**Supp. Fig. 1**). ConvNext models are modernized versions of Residual neural networks^15,16^ and include a “patchify” stem, GELU activation functions, significantly smaller initial model weights, and the use of stochastic depth, among other architectural and training improvements. For all models, the input consisted solely of the ECG waveform, with no demographic, MUSE-computed ECG measurements, or other features.

We select ConvNext hyperparameters by choosing those which produce the lowest loss during hyperparameter tuning for the initial VICReg pre-training detailed below. We hyperparameter tune over the: dimension of the convolution layers, rate at which the dimension of the convolution layers increases per stage, convolutional kernel size, stage ratio (number of blocks per ‘stage’), number of convolutional layers per block, patchify kernel size, initial value for the layer scaling, standard deviation for the layer weight initialization, stochastic depth rate, and average or max pooling use in the global pooling layer. For both the VICReg and Standard models, we use an initial layer dimension of 16 with a scale factor of 1.5 per stage; 4 stages of 3, 3, 27 and 3 blocks; 4 convolutional layers per block; convolutional kernel size of 5; a patchify kernel size of 12; an initial layer scaling value of 1e-05; a standard deviation of 0.1 for the layer weight initialization; a stochastic depth rate of 0.2; and average pooling.

During the VICReg pre-training detailed below, the global pooling is followed by an ‘expander’ head. The expander head is three dense layers, each of which is four times the size of the global pooling layer, with batch norm and ReLu in between them. When the VICReg baseline model is finetuned, this ‘expander’ head is removed, and replaced by a multilayer perceptron (MLP) that reduces the dimensionality of the output to 1. During training of the Standard Model, we test both the ConvNext structure with a MLP model head, and with an expander head, followed by a standard head, and find the latter produces better results on the validation set.

### VICReg Pre-Training

We used the VICReg^5^ method with the aforementioned ConvNext model structure for self-supervised representation learning (**Fig. 3**). In the VICReg method, the Variance term is designed to assure the model gives samples sufficiently distinct values for each given variable in the embedding. The Invariance term is designed to push the embeddings for augmented copies of a sample to be similar. The Covariance term is designed to assure that the model creates informational embeddings, by penalizing it for encoding correlated information in pairs of variables in the embedding. Specifically, the model is trained to minimize the loss function:

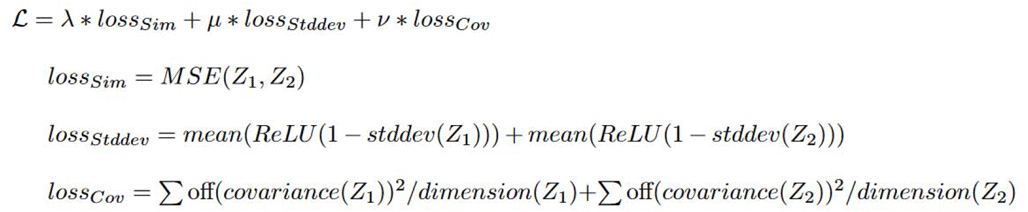

where lambda, mu, and nu are constants; Z_1_ and Z_2_ are the two matrices (batches of vectors) output by the model for two augmented copies of a batch of samples; off(covariance(Z)) is the non-diagonal components of the covariance matrix of Z, and dimension(Z) is non-batch dimension of the output matrix. As in the VICReg publication^5^, we use a Siamese net architecture, wherein both branches are the same model with the shared weights, so it is only one model producing both embedding outputs. For data augmentation, we randomly ‘shift’ the starting point of the 10-second ECG to any time point and copy the data that precedes the new start onto the end of the ECG, so it remains 10-seconds. After shifting, we permute the ECG using equally sized pieces, then scale the entire ECG by a value randomly sampled from a Gaussian distribution with a mean of one^17^. Lastly, we add Gaussian noise with a mean of zero to each sample point in the waveform from each lead. We include the number of the permutations, as well as the standard deviations of the scaling factor and of the noise to the hyperparameters used during the VICReg pre-training. We further include the scaling factors for the variance, invariance, and covariance loss (although we quickly settle on the values proposed in the original work^5^), the learning rate, and whether to use the ADAM or SGD optimizer in addition to the previously mentioned ConvNext hyperparameters.

### Deep learning model testing

After pre-training, the VICReg baseline model was fine-tuned to classify type 1 BrS ECG patterns on the BrS Training set and hereafter called VICReg model. A model with equivalent architecture but without the VICReg pre-training, hereafter called Standard model, was also trained on the same training set to classify BrS ECG patterns (**Fig. 1**). In both cases, we used 10-fold cross-validation on the training and validation sets comprising the BrS Training set. For each collection of hyperparameters tested, on each of the 10 folds a new model instance is created with randomly initialized weights for the Standard model, or with the VICReg baseline model weights for the VICReg model. Due to the instability of the Standard model’s training, we saved only instances that achieved an AUROC of at least 0.85 on their given training and validation folds, and we treated all copies of a model saved across the 10 folds as elements in an ensemble model to improve generalization. We also allowed for one re-initialization of training on each fold if the model produces a train or validation AUROC of less than 0.5 on that fold. If either the train or validation AUC on the fold was less than 0.85, the model instance was not saved, and thus was not incorporated into the model ensemble constructed for that set of hyperparameters. To finalize model selection, we re-tested the ensembles on the training/validation data and selected the VICReg and Standard model ensembles with the highest AUROC across the training and validation sets.

### Statistical Analysis

When comparing diagnosis prevalence between BrS and non-BrS cases, we compute odds ratios and provide the associated 95% confidence intervals (CIs). *P* Values were computed using Barnard’s Test. Values of *p* <0.05 and by extent 95% CIs that do not contain the value 1, were considered statistically significant. For BrS population prevalence estimates, we compute the confidence intervals with bias-corrected and accelerated bootstrapping using 10,000 resamples. Statistical analysis was performed using the Python libraries SciPy version 1.10.1, Scikit-learn version 1.2.2, and NumPy version 1.23.5. All model training and evaluation was performed using TensorFlow Version 2.3 and Python version 3.7.

## Acknowledgments

None

## Author contributions

RR performed data extraction, collection and analysis, designed and trained the models and contributed to data interpretation. CT contributed to study design, data interpretation and manuscript preparation. LC and LJ were the principal investigators and contributed to the study idea and design, interpreted the data, and revised the manuscript.

## Data availability

The data collected from NYU Langone Health during this study are patient data obtained under the Institutional Review Board’s ethical approval. The authors agree to share de-identified individual participant data and the study protocol with academic researchers following completion of a data use agreement specifying that this information cannot be shared. The coding used to train the AI model is dependent on annotation, infrastructure, and hardware and therefore, cannot be released.

---

**Supplemental Figure 1:**
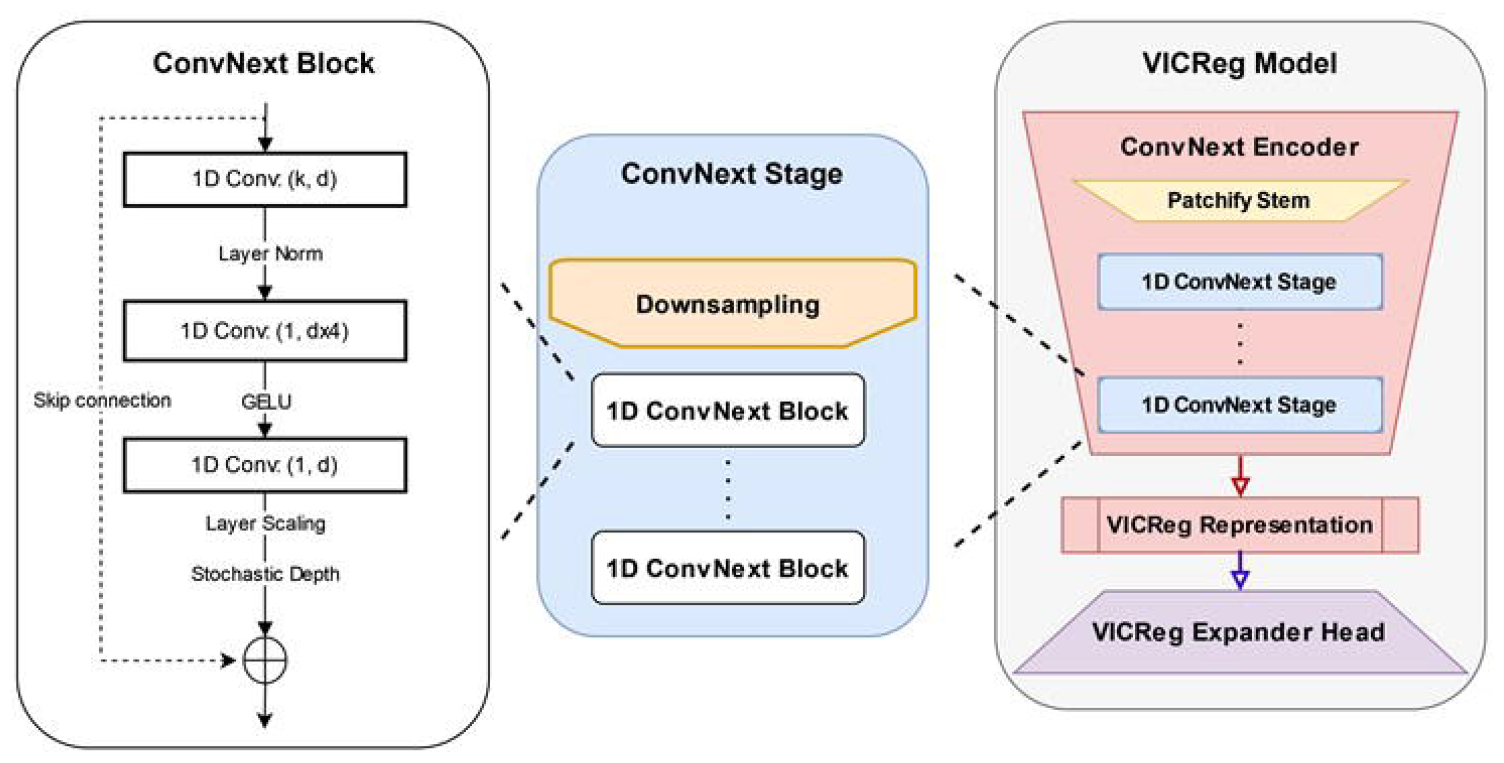
Model architecture of the ConvNext CNN used for both the Standard and VICReg models.

## Bibliography

1. Siontis, K.C., Noseworthy, P.A., Attia, Z.I. & Friedman, P.A. Artificial intelligence-enhanced electrocardiography in cardiovascular disease management. Nat. Rev. Cardiol. 18, 465–478 (2021).

2. Hannun, A.Y. et al. Cardiologist-level arrhythmia detection and classification in ambulatory electrocardiograms using a deep neural network. Nat. Med. 25, 65–69 (2019).

3. Ribeiro, A. H. et al. Automatic diagnosis of the 12-lead ECG using a deep neural network. Nat. Commun. 11, 1760 (2020).

4. Krahn, A.D. et al. Brugada Syndrome. JACC Clin. Electrophysiol. 8, 386–405 (2022).

5. Bardes, A., Ponce, J. & LeCun, Y. VICReg: Variance-Invariance-Covariance Regularization for Self-Supervised Learning. (2021).

6. Viskin, S. et al. Inaccurate electrocardiographic interpretation of long QT: The majority of physicians cannot recognize a long QT when they see one. Hear. Rhythm 2, 569–574 (2005).

7. Akdemir, I. Intermittent Brugada syndrome misdiagnosed as acute myocardial infarction and unmasked with propafenone. Heart 87, 543–543 (2002).

8. Rodríguez-Mañero, M. et al. T-Wave Oversensing in Patients With Brugada Syndrome: True Bipolar Versus Integrated Bipolar Implantable Cardioverter Defibrillator Leads. Circ. Arrhythmia Electrophysiol. 8, 792–798 (2015).

9. Liu, C.M. et al. A Deep Learning–Enabled Electrocardiogram Model for the Identification of a Rare Inherited Arrhythmia: Brugada Syndrome. Can. J. Cardiol. 38, 152–159 (2022).

10. Melo, L. et al. Deep learning unmasks the ECG signature of Brugada syndrome. PNAS Nexus 2, (2023).

11. Liao, S. et al. Use of Wearable Technology and Deep Learning to Improve the Diagnosis of Brugada Syndrome. JACC Clin. Electrophysiol. 8, 1010–1020 (2022).

12. Patel, S.S., Anees, S. & Ferrick, K.J. Prevalence of a Brugada pattern electrocardiogram in an urban population in the United States. Pacing Clin. Electrophysiol. 32, 704–8 (2009).

13. Vutthikraivit, W. et al. Worldwide Prevalence of Brugada Syndrome: A Systematic Review and Meta-Analysis. Acta Cardiol. Sin. 34, 267–277 (2018).

14. Liu, Z. et al. A ConvNet for the 2020s. (2022).

15. He, K., Zhang, X., Ren, S. & Sun, J. Deep Residual Learning for Image Recognition. In 2016 IEEE Conference on Computer Vision and Pattern Recognition (CVPR) 770–778 (IEEE, 2016). doi:10.1109/CVPR.2016.90.

16. He, K., Zhang, X., Ren, S. & Sun, J. Identity Mappings in Deep Residual Networks. in 630–645 (2016). doi:10.1007/978-3-319-46493-0_38.

17. Iwana, B.K. & Uchida, S. An Empirical Survey of Data Augmentation for Time Series Classification with Neural Networks. (2020) doi:10.1371/journal.pone.0254841.

